## Supplementary for "Collecting genetic samples and linked mental health data from adolescents in schools: Protocol co-production and a mixed-methods pilot of feasibility and acceptability"

**Supplementary Table 1. MAGES teacher focus group schedule**

| **Mental health research in young people** |
|---|
| Do you think schools should be involved in this kind of research?<br>*PROMPT Do you think your school should be involved in research on mental wellbeing and genetics?*<br>*PROMPT How did you feel about your school taking part in research on mental wellbeing and genetics?* |
| What do you think of researchers collecting DNA samples from children in school during the school day facilitated by teaching staff? |
| What do you think of linking child genetic data to other records? |
| **Evaluation of MAGES (practicalities)** |
| How did you find being a part of MAGES in your school?<br>*PROMPT What were the good parts?*<br>*Was it beneficial?*<br>*PROMPT What were the bad parts?*<br>*Did it increase your workload significantly?*<br>*What was most time consuming?*<br>*PROMPT How could we improve?* |
| How did you/ would you 'sell' this study to parents? To students? |
| Do you think the way the study was run (i.e. the study design) was practical?<br>*PROMPT What practical changes would you make to make it easier for schools to take part?* |
| What would be the best way to get staff within your school to engage with MAGES? |
| **Evaluation of the school/parents/pupils** |
| How does your school communicate with parents? |
| Did any parents approach you with concerns about MAGES?<br>*PROMPT For instance, were there any concerns about taking DNA samples from children?* |
| Did students approach you with any concerns before/ after workshop/ saliva collection? |
| Did any parents approach you for any further information about MAGES?<br>*PROMPT Did you know we have a website?* |
| **Beyond the current MAGES** |
| Do you think this would work on a larger scale?<br>*PROMPT How about practicalities? Do you think teachers would be interested in delivering the science workshops themselves?*<br>*PROMPT What things (incentives) would encourage all schools in Wales to take part?* |
| Could you see your school participating in research like this in future?<br>*PROMPT Knowing what you do now, would you take part again?*<br>*PROMPT How likely would you be to take part again and why?* |

**<u>Supplementary Figure 1. Sticker chart used for student feedback following each science workshop</u>**

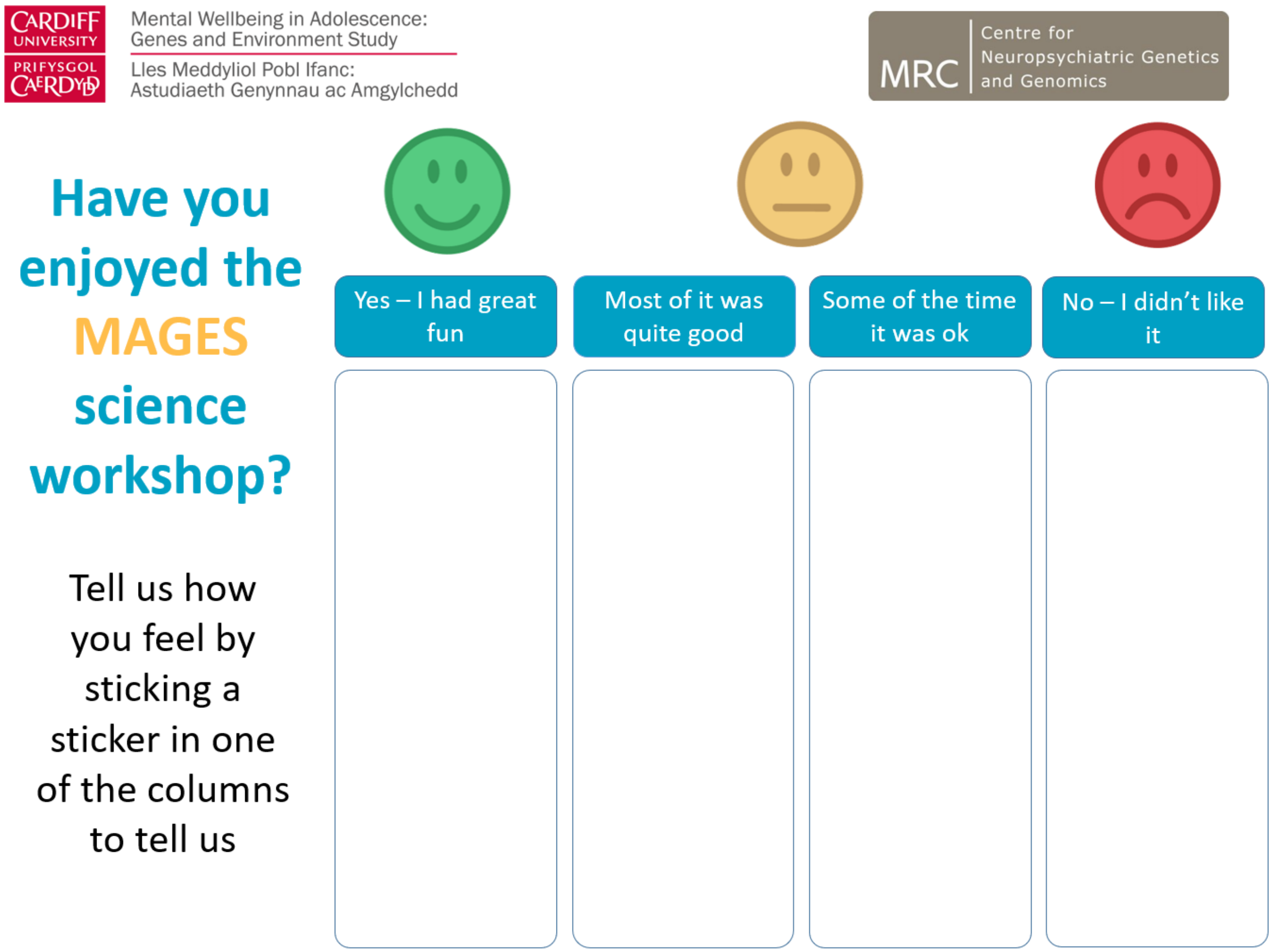

**Supplementary Figure 2. Student instructions for providing a saliva sample**

### How to give a spit sample

There is one saliva (spit) pot for you to fill. It's quick and easy.

Please don't eat, drink, smoke or chew gum for 30 minutes before giving your spit sample. Also, please don't remove the plastic film from the lid.

You can then follow these simple instructions:

1. Spit saliva into the empty container, until it reaches the fill line shown below (not including bubbles):

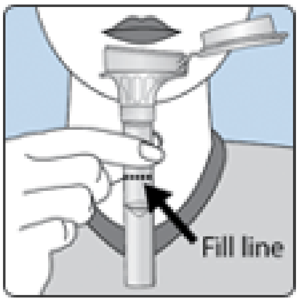


2. Hold the tube upright with one hand. Close the funnel lid with the other hand (as shown) by firmly pushing the lid until you hear a **loud click**. The liquid in the lid will be released into the tube to mix with the saliva. Make sure that the lid is closed tightly.

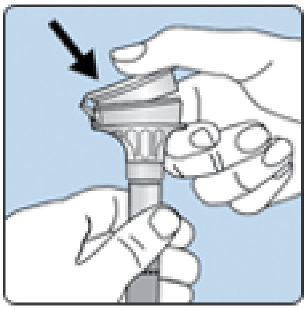

3. Hold the tube upright. Unscrew the funnel from the tube.

4. Use the small cap to close the tube tightly.

5. Shake the capped tube for 5 seconds. You can discard or recycle the funnel.

6. Place into the plastic box provided and seal.

7. Return the filled tube to the researcher.
